# Computerized physical and cognitive training improves the functional architecture of the brain in adults with Down Syndrome: a network science EEG study

**DOI:** 10.1101/2020.05.28.20115709

**Authors:** Alexandra Anagnostopoulou, Charis Styliadis, Panagiotis Kartsidis, Evangelia Romanopoulou, Vasiliki Zilidou, Chrysi Karali, Maria Karagianni, Manousos Klados, Evangelos Paraskevopoulos, Panagiotis D. Bamidis

## Abstract

Understanding the neuroplastic capacity of people with Down Syndrome (PwDS) can potentially reveal the causal relationship between aberrant brain organization and phenotypic characteristics. We used resting-state EEG recordings to identify how a neuroplasticity-triggering training protocol relates to changes in the functional connectivity of the brain’s intrinsic cortical networks. Brain activity of 12 PwDS before and after a ten-week protocol of combined physical and cognitive training was statistically compared to quantify changes in directed functional connectivity in conjunction with psychosomatometric assessments. PwDS showed increased connectivity within the left hemisphere and from left to right hemisphere, as well as increased physical and cognitive performance. Our findings reveal a strong adaptive neuroplastic reorganization as a result of the training that leads to a less-random network with a more pronounced hierarchical organization. Our results go beyond previous findings by indicating a transition to a healthier, more efficient, and flexible network architecture, with improved integration and segregation abilities in the brain of PwDS. Resting-state electrophysiological brain activity is used here for the first time to display meaningful relationships to underlying DS processes and outcomes of importance in a translational inquiry. This trial is registered with ClinicalTrials.gov Identifier NCT04390321.

**Author Summary:** The effects of cognitive and physical training on the neuroplasticity attributes of people with and without cognitive impairment have been well documented via neurophysiological evaluations and network science indices. However, there is still insufficient evidence for people with Down Syndrome (PwDS). We investigated the effects of a combinational training protocol on the brain network organization of 12 adult PwDS using EEG and network indices coupled with tests assessing their cognitive and physical capacity. We report evidence of adaptational neuroplastic effects, pointing to a transitional state towards a healthier organization with an increased ability to integrate and segregate information. Our findings underline the ability of the DS brain to respond to the cognitive demands of external stimuli, reflecting the possibility of developing independent-living skills.

## Introduction

Neuroplasticity can emerge in both typical and atypical brains and allows for either development, reaction, recovery, or adaptation to internal and external stimuli (Trojan & Pokorny, 1999). In contrast to typically developed (TD) individuals, the Down Syndrome (DS) brain presents atypical levels of inhibition due to gene over-expression, leading to prolonged failure of synaptic plasticity and a reduced capacity for remodeling (Baroncelli et al., 2011). These characteristics, along with morphogenetic modifications, are among the leading causes of brain disability in people with DS (PwDS) (Dierssen, Herault, & Estivill, 2009).

DS individuals, in comparison to age-matched TD individuals, have smaller frontal, amygdala, and cerebellar volumes (but increased parahippocampal volume) (Pinter, Eliez, Schmitt, Capone, & Reiss, 2001; White, Alkire, & Haier, 2003), a pattern which overpronounces after their 50^th^ year of age (Dierssen, 2012). The current consensus has associated the cognitive capabilities and deficiencies of PwDS with several distinct brain regions emphasizing an abnormal and less efficient DS brain organization (Menghini, Costanzo, & Vicari, 2011).

Electroencephalography (EEG) and magnetoencephalography (MEG) studies have complemented the notion of this atypical organization. PwDS, when compared to TD controls, exhibit slow brain wave, especially in left posterior areas (Babiloni et al., 2010, 2009), with higher delta band and lower alpha and beta band activity, a pattern also evident in patients with Alzheimer’s Disease (AD) (Babiloni et al., 2010, 2009; García-Alba et al., 2019; Hemmati, Ahmadlou, Gharib, Vameghi, & Sajedi, 2013). fMRI studies have shown a rather-simplified DS network organization (Anderson et al., 2013), lacking the appropriate efficiency and flexibility (Edgin, Clark, Massand, & Karmiloff-Smith, 2015). Given the limited capacity of the DS brain to consolidate information due to its disorganized architecture of reduced segregation and impaired integration, diffused connectivity (hyper-synchrony) (Anderson et al., 2013), and decreased long-range connectivity, the DS brain network’s potential for plasticity is in question (Edgin et al., 2015).

Nevertheless, the DS brain does possess neuroplastic capabilities, at least in the form of compensatory events (Haier, Head, Head, & Lott, 2008; Takashima et al., 2014). The emergence of AD-related characteristics (i.e., altered theta band activity) in PwDS reaffirms the possibility of compensatory mechanisms before and during the expression of AD (Takashima et al., 2014). Characterizing neuroplasticity in PwDS is vital in understanding causality between aberrant brain circuitry and the cognitive and behavioral phenotype. This step would allow the quantification of the remodeling potential of evidence-based interventions.

Interventional approaches have shifted from a pharmaceutical concept, with, so far, inconclusive results regarding beneficial effects (Costa, 2011; Kálmán et al., 2009; Kishnani et al., 2010; Lott et al., 2011; Mohan, Carpenter, & Bennett, 2009), to non-pharmaceutical interventions of physical and behavioral components, aiming to trigger neuroplasticity and enhance brain health or to protect against neurodegenerative events (Head et al., 2007). Such interventions have shown promise in healthy aging and populations with cognitive impairments and have provided mounting evidence for lifelong brain plasticity (Bamidis et al., 2014; Colcombe et al., 2006, 2004; Cotman, Berchtold, & Christie, 2007; Erickson et al., 2011; Klados, Styliadis, Frantzidis, Paraskevopoulos, & Bamidis, 2016; Kramer et al., 1999; Styliadis, Kartsidis, & Paraskevopoulos, 2015; Styliadis, Kartsidis, Paraskevopoulos, Ioannides, & Bamidis, 2015).

Despite evidence supporting the positive impact of exercise-based (Bertapelli, Pitetti, Agiovlasitis, & Guerra-Junior, 2016; Hardee & Fetters, 2017), as well as behavioral and cognitive interventions (Fonseca, Navatta, Bottino, & Miotto, 2015; McGlinchey, McCarron, Holland, & McCallion, 2019) for PwDS, DS literature features a substantial knowledge gap, as there are no reports of neurophysiological investigations of the training- induced effects, and a network-theory based assessment of the training-induced plasticity. Deviations in the brain functionality of PwDS have been previously addressed with the investigation of resting-state networks (RSNs) (Ahmadlou, Gharib, Hemmati, Vameghi, & Sajedi, 2013; Anderson et al., 2013; Babiloni et al., 2010, 2009; Pujol et al., 2015; Vega, Hohman, Pryweller, Dykens, & Thornton-Wells, 2015).

We hypothesized that combined physical (PT) and cognitive training (CT) would trigger neuroplasticity and reorganize the DS brain network to adapt to the increased cognitive and physical demand. Resting-state EEG data were acquired to characterize plasticity in the DS brain through network science and investigate how the training-induced neuroplasticity affects the organization and characteristics of the DS network, as opposed to the reported random-like architecture (Ahmadlou et al., 2013) with impaired integration and segregation capabilities.

Source analysis was performed using low resolution electromagnetic tomography (LORETA) (Pascual-Marqui, Michel, & Lehmann, 1994), and directed functional connectivity was estimated via phase transfer entropy (PTE) in 0.53-35 Hz. This measure quantifies the direction of the information flow, allowing for a whole-head analysis without requiring an *a priori* head model definition. A non-parametric permutation test was applied to extract the significant, within-network differences (post- vs. pre-intervention) in connectivity. A graph-theoretical analysis was performed to index the training’s neuroplastic effects through the statistical comparison of graph measures between the two time-points.

## Results

### Psychometric and somatometric results

Psychosomatometric score comparisons between the two time-points were performed with the use of non-parametric Wilcoxon tests and paired t-tests. Physical assessments exhibited a significant improvement in score for the arm curl test (Table 1). Significant differences in the time of completion (decrease in duration) were evident in the Time Up and Go assessment test (Table 1). The comparison of psychometric assessments revealed an increase in the Digits Forward score (Digits Span test), the Mazes test, as well as the Ravens AB and Ravens Total Score (Table 1). The rest of the tests showed no significant post-pre score changes.

**Table 1.** Somatometric and psychometric tests with significant differences post vs. pre, along with the post-pre mean difference. Results were considered significant for *p < 0.05*.

| Somatometric | p-value | mean difference |
| --- | --- | --- |
| Arm Curl | 0.017 | 1.75 |
| Time Up and Go | 0.007 | -1.67 |
| **Psychometric** |  |  |
| Digits Span (Digit Forward) | 0.032 | 1.50 |
| Mazes | 0.032 | 0.97 |
| Ravens AB | 0.017 | 2.12 |
| Ravens Total Score | 0.015 | 3.13 |

### PTE results

Statistical comparisons of the PTE matrices between the pre- and post-intervention EEG measurements indicate the reorganization of cortical connections (62 nodes and 40 edges), due to training, in the DS resting-state network (*p < 0.05*, corrected for multiple comparisons via the non-parametric NBS method (Zalesky, Fornito, & Bullmore, 2010), 5000 permutations) (Figure 1). Specifically, the cortical reorganization in the DS brain is characterized by the strengthening of connections within: i) the left hemisphere, from nodes in the occipital (cuneus and lingual gyrus) and the temporal lobe (fusiform gyrus) to the frontal lobe (superior and middle gyrus) and from the frontal lobe to the parietal lobe (superior, precuneus and postcentral gyrus), ii) the right hemisphere, from nodes in the occipital (cuneus, lingual and middle gyrus) and temporal (fusiform and inferior gyrus) lobe to the frontal lobe (superior, cingulate and medial gyrus), and between: i) nodes of the left fusiform gyrus, and inferior temporal lobe and the right frontal lobe (superior, middle and inferior gyrus), ii) the right middle occipital lobe, sub-gyral, parahippocampal and fusiform gyrus to the left frontal lobe (superior, middle, precentral gyrus), iii) the left inferior temporal lobe to the right anterior cingulate, and iv) the right frontal lobe (middle and inferior gyrus) to the left postcentral and middle frontal gyrus (Figure 1). In respect to the core RSNs, as defined by Yeo et al. (Yeo et al., 2011), we have noted increased connectivity within the dorsal attention network (DAN), and between the visual (VIS) and frontoparietal network (FPN), VIS and default-mode-network (DMN), VIS and ventral attention network (VAN), VAN and FPN, FPN and DMN, FPN and somatosensory network (SMN), FPN and DAN, DAN and DMN, as well as DAN and SMN (Table 2).

**Figure 1.**
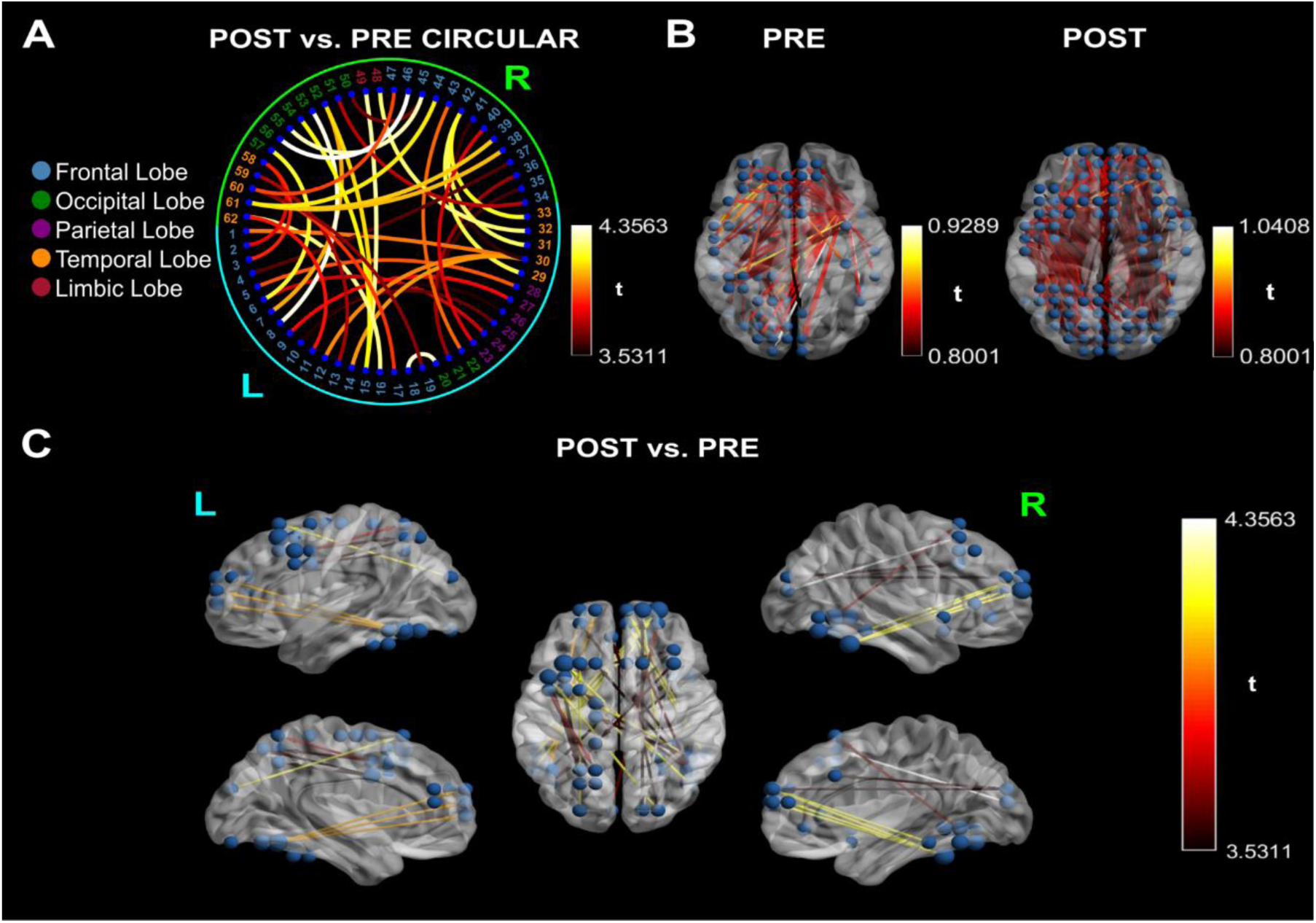
Cortical connectivity between post- and pre-intervention networks and for each time point (t-value>3.52). The color scales represent t-values. **A:** Circular graph depicting the cortical reorganization in the DS brain (cyan: left hemisphere, lime: right hemisphere, blue: frontal lobe, green: occipital lobe, purple: parietal lobe, orange: temporal lobe, maroon: limbic lobe). The cortical reorganization is characterized by the strengthening of direct connections within: i) the left hemisphere, from nodes in the occipital and temporal lobe to the frontal lobe, and from the frontal lobe to the parietal lobe, ii) the right hemisphere, from nodes in the occipital and temporal lobe to the frontal lobe, and between: i) nodes of the left fusiform gyrus, and inferior temporal lobe and the right frontal lobe, ii) the right middle occipital lobe, sub-gyral, parahippocampal and fusiform gyrus to the left frontal lobe, iii) the left inferior temporal lobe to the right anterior cingulate, and iv) the right frontal lobe to the left postcentral and middle frontal gyrus. **B:** Depiction of pre- and post-intervention networks in comparison to the null hypothesis. The t-values indicate that the post-resting-state network has significantly stronger connections than the pre network. **C: Post vs. Pre.** Significant post-pre connectivity differences. Information direction is depicted through line arrows. The visualized networks are significant at a level of *p < 0.05*, corrected for multiple comparisons via the non-parametric NBS method with an internal threshold of 0.8. The difference in nodal size depicts the increase in the node degree centrality; the nodes with the most increased connectivity are located at the left parietal lobe.

**Table 2.**
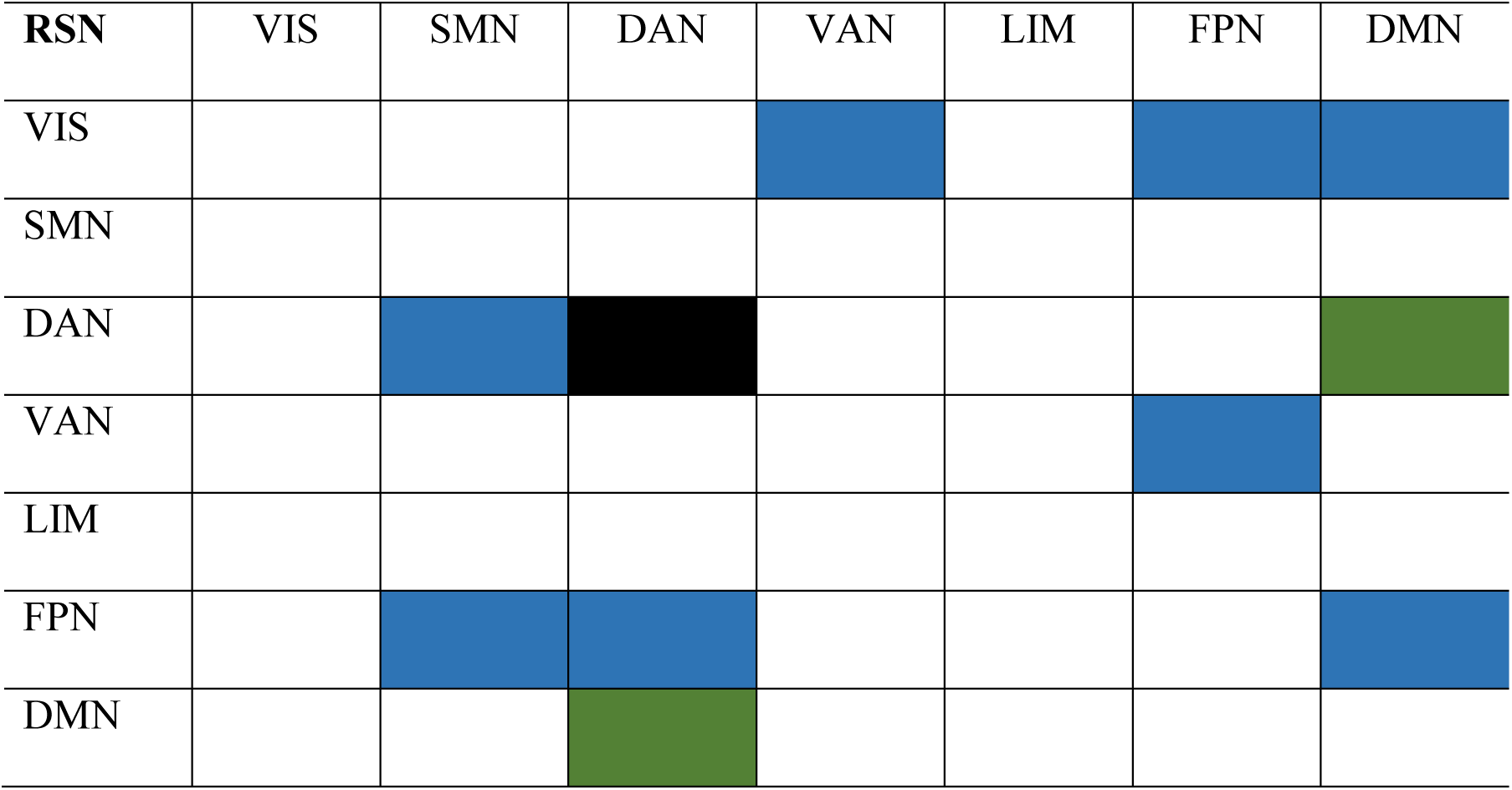
Significant increase in connectivity of RSNs as defined by Yeo et al. (Yeo et al., 2011). The black color signifies the within-network connectivity, the green color the bilateral connectivity between-networks, and the blue color the unilateral connectivity between-networks from the RSN in the first column to the RSN in the first row.

### Graph measures

The significant changes (*p < 0.05*) in global efficiency (GE), transitivity (TS), characteristic path length (CPL), clustering coefficient (CC), betweenness centrality (BC), and the small-worldness (SW) measure, sigma (*σ*), between the pre- and post-networks were investigated using Analysis of Covariance, where graph measures served as the dependent values and density as the covariate (Table 3). Regarding the global measures, we report increases in GE and TS and a decrease in CPL. From the comparison of local measures, CC shows significant differences in 716 nodes, with increased values in 607 of these nodes (Figure 2). From these, 323 nodes belong to the left hemisphere (102 in the frontal lobe (FL), 39 in the central midline (CM), 56 in the parietal lobe (PL), 49 in the temporal lobe (TL), 26 in the limbic lobe (LL), 49 in the occipital lobe (OL)) and 284 to the right hemisphere (76 in FL, 51 in CM, 37 in PL, 49 in TL, 28 in LL, 44 in OL) (Figure 2). From the significant nodes, 109 showcased a significant decrease in value, with 48 nodes localized in the left hemisphere (7 in FL, 6 in CM, 6 in PL, 19 in TL, 2 in LL, 8 in OL) and 61 in the right hemisphere (22 in FL, 2 in CM, 8 in PL, 9 in TL, 3 in LL, 17 in OL). BC increased in 15 nodes with 9 pertaining to the left hemisphere (4 in the medial FL, 2 in PL (precuneus and superior PL), 1 in the inferior TL, 1 in the cingulate gyrus of LL, 1 in the middle OL) and 6 to the right (2 in FL (inferior and medial gyrus), 2 in the precuneus (PL), 1 in the cingulate gyrus (LL), 1 in the lingual gyrus (OL)), while it decreased in 2 nodes of the right angular gyrus (LL). The post vs. pre comparison of the SW measure revealed a significant increase in the σ-value, but the values for both the pre- and post-intervention networks did not exceed the SW threshold of *σ*>1. We performed a multi-scale Louvain community detection, which showed that the networks of the two time-points are organized in different levels of hierarchy. The community analysis on the pre-intervention DS network indicated that its communities are nested within 3 levels of hierarchy with the number of communities across all levels ranging from 70 communities to 442, while the communities of the post DS network are organized in 4 levels, with their number ranging from 45 to 290 across all levels.

**Figure 2.**
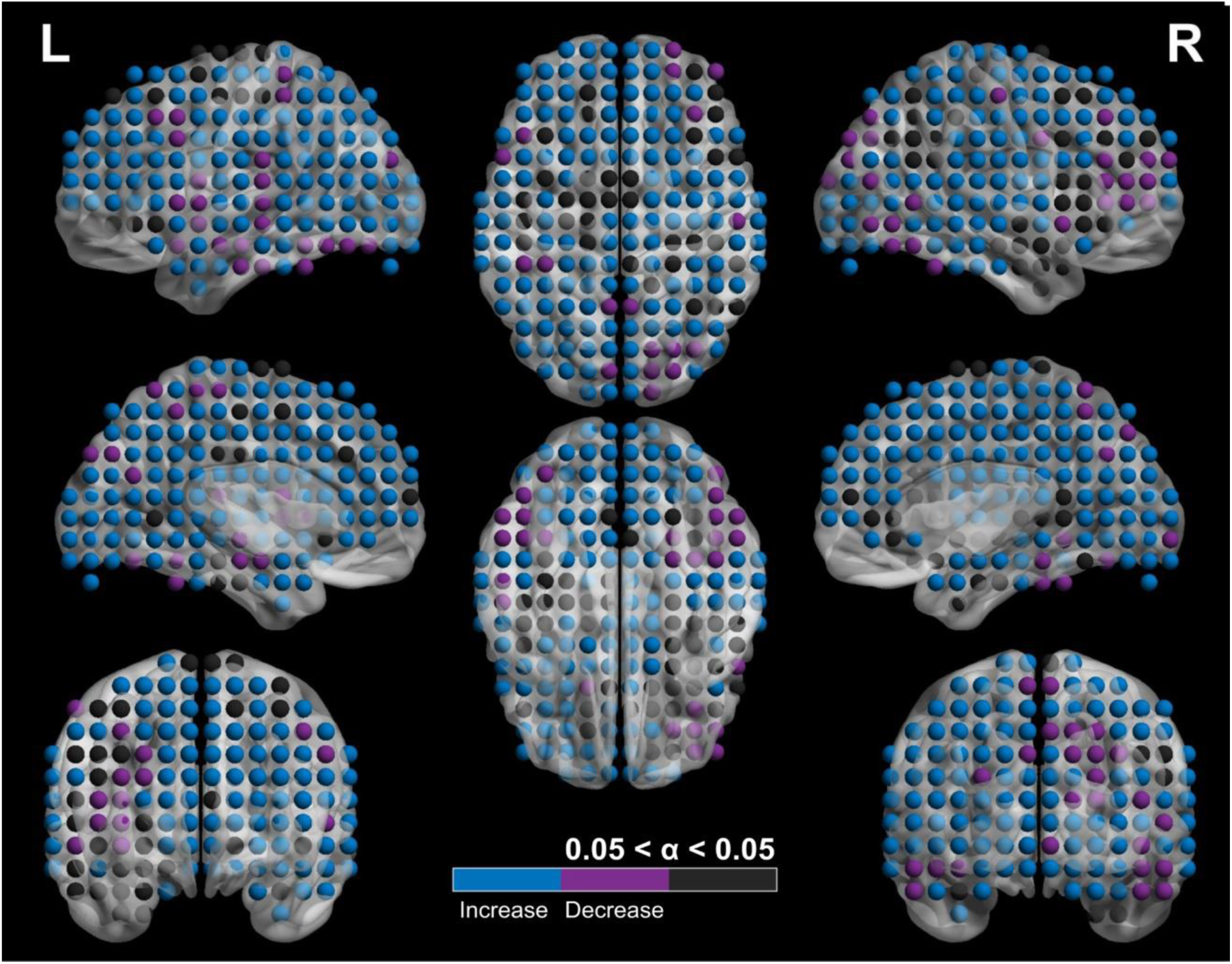
Depiction of the shifts in the CC of nodes. The colormap indicates significance at an alpha-value (*a < 0.05*) coupled with the direction of the shift. The black nodes have no significant shifts in CC values post- and pre-intervention, blue nodes show a significant increase at *a < 0.05*, while magenta nodes show a significant decrease at *a < 0.05*.

**Table 3.**
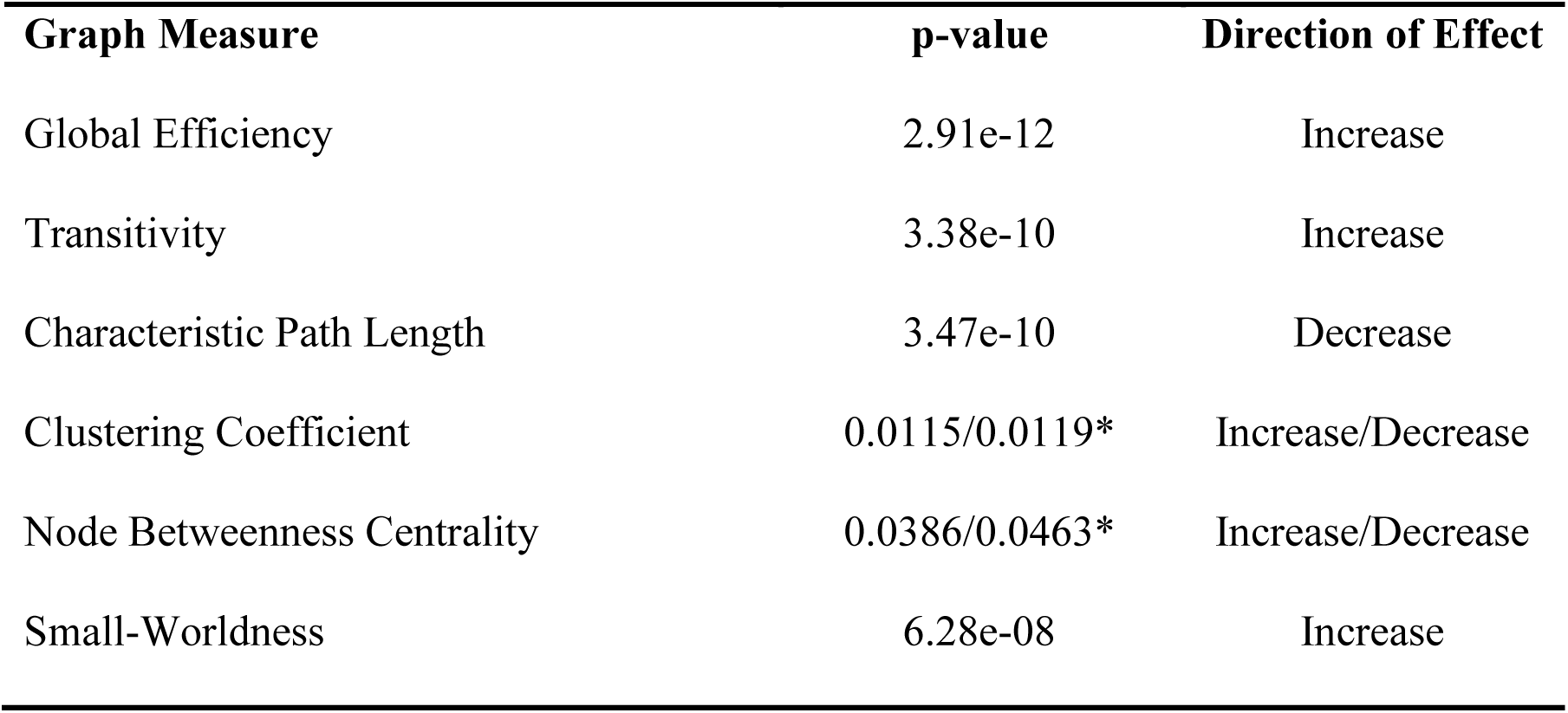
Significant post-pre changes in global (Global efficiency, Transitivity, Characteristic Path Length) and local (Local Clustering Coefficient, Node Betweenness Centrality) graph measures as well as the small-worldness measure. Results were considered significant for *p < 0.05*. *For the local graph measures, the reported p-value is the mean value of the nodes showing a significant increase and a significant decrease, respectively.

## Discussion

A ten-week intervention of combined PT and CT in adults with DS can trigger neuroplasticity resulting in cortical reorganization, quantified via EEG indices, and graph measures. Our results reveal that: i) short-term training can modify the physical and cognitive performance of adults with DS, ii) the DS brain can adapt to novel stimulation and challenges by utilizing neuroplasticity and reorganizing itself, and iii) adaptive neuroplasticity may lead to a more flexible and healthier functional network.

### Short-term training modifies physical and cognitive performance

Our PT findings indicate the significant improvement of physical capacity for upper body strength and endurance, mobility, static and dynamic balance, and are in line with previous findings on adult PwDS (Hardee & Fetters, 2017; Ruiz-González, Lucena-Antón, Salazar, Martín-Valero, & Moral-Munoz, 2019). However, our protocol targets multiple physical domains, while most studies investigating the effect of PT on the somatic capacity of adult PwDS (Hardee & Fetters, 2017; Ruiz-González et al., 2019) have mainly focused on a specific domain, i.e. resistance training or aerobic training and, less often, on a combination of two or more domains.

With respect to CT, our PwDS exhibited an improved general cognitive capacity, i.e., in the level of general intelligence, planning and organization skills, and in short-term memory, attention, and concentration. Previous cognitive interventions on adult PwDS have mainly focused on memory (Fonseca et al., 2015) and, less often, on executive functions (McGlinchey et al., 2019). In line with the findings of the latter study, adult PwDS can complete a computerized CT program that targets multiple domains of cognition and exhibit significant improvements.

### Cortical reorganization in the DS brain and neuroplastic capacity

PwDS present capacity for neuroplasticity. Regarding the type of neuroplasticity that is expressed here, we can rule out the triggering of evolutionary and reactive plasticity since the first is most active during the development of the brain (Trojan & Pokorny, 1999), and the latter is most commonly triggered by a single stimulus (Trojan & Pokorny, 1999) (our training utilizes repetitive stimuli). Adaptational and reparation plasticity can both cause cortical reorganization through long-term stimulation. Here, combined PT and CT have increased the communication within and between nodes of the two hemispheres (Figure 1A&C). The DS cortical networks are characterized by a simplified organization (Anderson et al., 2013) with hyper-synchrony between brain regions (Anderson et al., 2013; Edgin et al., 2015; Vega et al., 2015). Therefore, the reported shifts in connectivity indicate the emergence of a reorganized network, which can be the result of either adaptational or reparation plasticity. Our finding regarding the increased left intra-hemispheric information flow can be regarded as the result of adaptational neuroplasticity being triggered in the DS brain given the atypical right-hemispheric functional preference (Elliott, Weeks, & Chua, 1994). However, the left-to-right hemisphere directionality is a pattern that has been previously evidenced in young PwDS (Babiloni et al., 2009), and its enhancement in adult PwDS may indicate that the training has triggered reparation neuroplasticity.

The enhanced connectivity within (DAN) and between (VIS and FPN, VIS and DMN, VIS and VAN, VAN and FPN, FPN and DMN, FPN and SMN, FPN and DAN, DAN and DMN, as well as DAN and SMN) the core RSNs (Yeo et al., 2011), further highlights the post-intervention DS functional reorganization. In respect to the aforementioned findings of Anderson and colleagues (Anderson et al., 2013), the strengthening of connections between distinct RSNs may also favor that the induced neuroplastic attributes represent adaptation. Given the evidence pointing towards an inverse relationship between the level of cognitive impairment of PwDS and the within DAN (Pujol et al., 2015; Vega et al., 2015) connectivity (i.e., the lower the DAN connectivity, the higher the cognitive impairment for DS), we suggest that the increased connectivity within DAN may associate with the increased cognitive capacity due to training in our DS sample. This would rule out the possibility that the type of plasticity represents reparation. In essence, future studies could potentially confirm or rule out this possibility.

The cortical reorganization, in conjunction with the increases in general intelligence, indicates that the DS brain has entered a more flexible state. The DS network has been previously reported to lack the flexibility that characterizes the TD brain (Edgin et al., 2015) and subsequently higher intelligence (Barbey, 2018; Hilger, Fukushima, Sporns, & Fiebach, 2019). Flexibility can reflect the brain’s capacity to adapt to novel stimulations (Barbey, 2018). It is plausible that the transition to a healthier organization emerges from the increased DS network flexibility and further points to the triggering of adaptational neuroplastic attributes.

### Graph theory characteristics support the neuroplastic transition towards a healthier DS network organization

We suggest that the network’s reorganization indexes a transitional state from a random-like network towards a healthier functional architecture, exhibiting less-random characteristics, increased flexibility and robustness, and improved integration and segregation capabilities (please see Figure 3 for an illustration of our theoretical proposal). In close relation, the SW measure, which classifies the pre- and post-intervention networks as non-SW, shows a significant increase in the post DS network and can, in turn, attribute the adult DS functional network (pre-intervention network) with features of a random network (Ahmadlou et al., 2013). Thus, the increased SW can index a transition to a less- random organization. The randomness of the pre-DS network is not surprising, given that basic cognitive abilities (low level of general intelligence, which also characterizes PwDS) are exemplified by random networks, while broad cognitive abilities (high level of general intelligence) would require a SW organization (Barbey, 2018; Hilger et al., 2019; Watts & Strogatz, 1998). SW networks maintain an optimal balance of integration and segregation (combining random and regular network characteristics) to support general and specific abilities (Barbey, 2018; Hilger et al., 2019; Watts & Strogatz, 1998). In contrast, the absence of SW properties in the DS brain indicates the increased risk of communication loss between connected regions and their causality for cognitive decline (Ahmadlou et al., 2013; Sanz et al., 2010).

**Figure 3.**
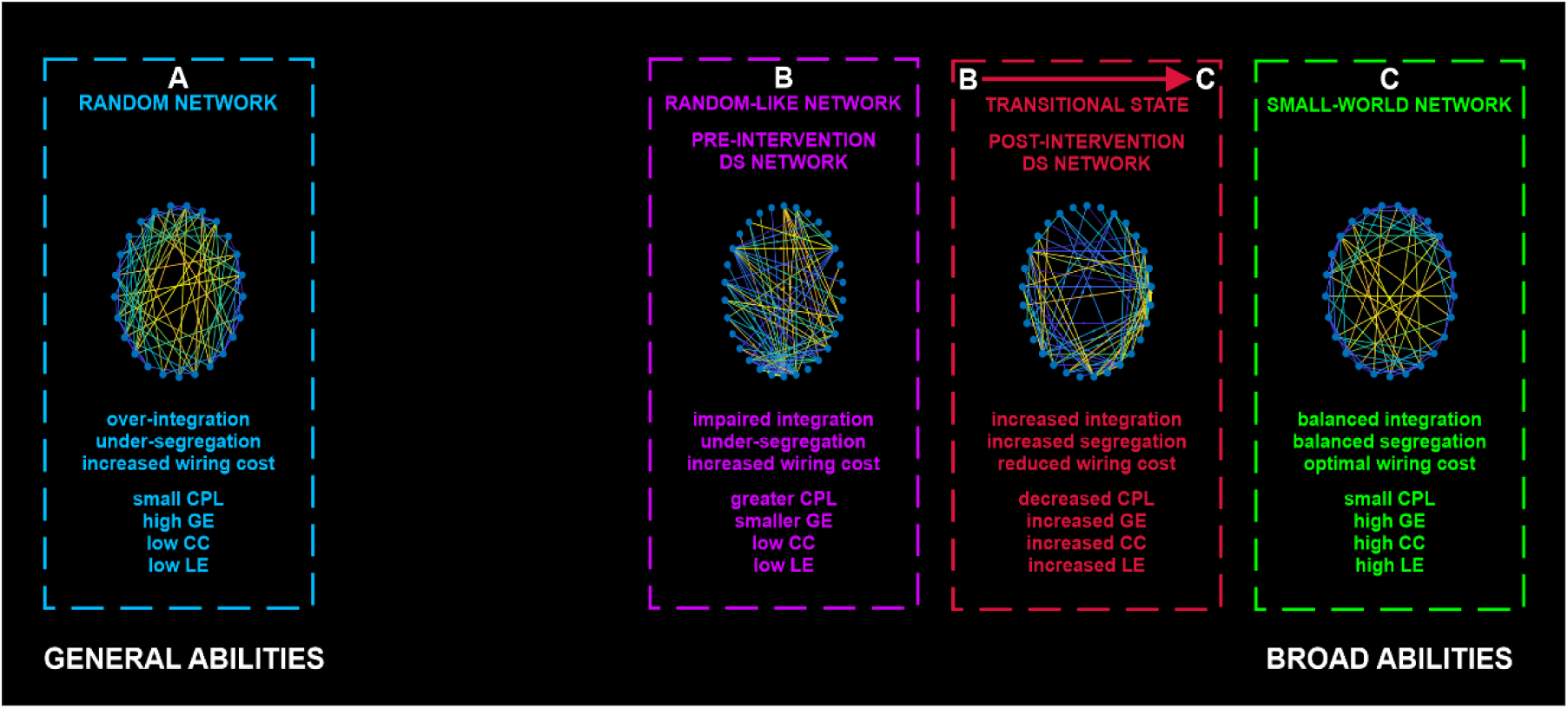
Illustration of our theoretical proposal. Changes in connectivity and graph-theory characteristics, as an outcome of the adaptational neuroplasticity in the DS brain, characterize the post-intervention DS network (**➔ C**) as a transitional state from the random-like organization of the pre-intervention DS network (Ahmadlou et al., 2013) (**Β**) towards a healthier functional structure. Random networks (Erdös–Rényi, random graph) (**Α**) are characterized by high global efficiency (over-integration) and low clustering (under-segregation) and exemplify the general abilities of general intelligence (low intelligence level). Small-world networks (**C**) feature the functional organization of TD brain networks and incorporate characteristics of both random and regular networks, achieving an optimal balance between global and local characteristics. SW networks (**C**) are associated with the broad abilities’ component of general intelligence (higher intelligence level). The pre-intervention DS network (**B**) showed a random-like, simplified architecture, with impaired segregation and integration, as evidenced by the low CC (random network characteristic) and decreased GE and increased CPL, respectively. This is in line with previous literature (Ahmadlou et al., 2013; Anderson et al., 2013; Pujol et al., 2015; Vega et al., 2015). The DS pre-network’s (**B**) integration-related characteristics (lower GE, higher CPL) are not common in random networks. Hence, the DS pre-network (**B**) is classified as random-like and not entirely random, maintaining an equal distance from random (**A**) and SW (**C**) networks. The DS post-network (**➔ C**) exhibits an increase in integration (random network characteristic), as well as segregation (regular network characteristic), so it is interpreted as a step towards an SW- like architecture that highlights a healthier brain organization.

Considering the DS brain (lack of functional specialization and impaired integration (Anderson et al., 2013), the simultaneous increment in GE and TS denotes the increase of both integration and segregation and the transition to a healthier network organization, with the rise in TS further supporting the reorganization to a less-random architecture. The decrease of CPL in TD individuals could be interpreted as a contradicting shift towards a more random network that disturbs the SW balance. In our case, the CPL is a step towards the opposite direction since the adult DS network deviates from random networks by displaying a greater CPL, thus characterizing it as “random-like” (Figure 3). Previous fMRI studies report that the brain network of PwDS presents decreased long-range connectivity (Anderson et al., 2013; Pujol et al., 2015) and thus increased CPL (Bassett & Bullmore, 2006). Hence, the decreased CPL supports our hypothesis and, coupled with the previously mentioned shifts, signals the decrease in wiring cost and subsequent increase in cost-efficacy (Achard & Bullmore, 2007; Bullmore & Sporns, 2012).

Our hypothesis of a shift towards a healthier network organization is also evident from a local network perspective. The increased CC in most areas of the brain indicates a rise in robustness, and fault tolerance in the network (Achard & Bullmore, 2007), which can potentially serve in neuroprotection for PwDS (Ahmadlou et al., 2013). Still, the effectiveness of such a mechanism would rely on the trade-off between integration and segregation, mainly maintaining SW network characteristics (Stam, Jones, Nolte, Breakspear, & Scheltens, 2007). The DS network, when compared to a TD network architecture of distributed communities, has been found to exhibit a lobar organization with increased local communication (Anderson et al., 2013) and reduced BC (Hemmati et al., 2013) making the aggregation of information less efficient. The aforementioned evidence, coupled with our increased hierarchical organization of communities in the post DS network, and the increase of BC in nodes, support the transition to a more hierarchical organization, rendering the network more efficient in the coherent distribution of information.

### Future directions and limitations

A limitation of the study was the small sample size of the experimental group. Our results could potentially be interpreted differently in reference to a control group (TD or passive DS). Though this study is the first to provide insights into the benefits of combined PT and CT in adults with DS through neuropsychological and neurophysiological assessments, a gap remains in our basic understanding of the sole contributions of each training to DS related cognition and brain function. Further studies are necessary to index the influence of each component on the triggering of neuroplasticity and subsequently network organization and cognition. Similarly, the stability of the reported neuroplastic shifts can only be addressed through additional and more in-depth investigations, including follow-up studies, to fill the gaps in the existing literature. Finally, the correlation of the somatometric and/or psychometric scores to the graph measures would increase the strength of our results, as it would indicate that the neuroplastic effect that we report is also reflected in behavior. We explored such a relationship between the changes in graph measures and the shifts in psychosomatic assessments (which showed post-pre significant changes) via a: i) correlation, ii) linear regression, and iii) multiple linear regression. The lack of a significant correlation was probably due to the small sample size, which did not produce enough power to quantify the effect, or due to the greater sensitivity that the neuroimaging methods show with regard to learning-induced neuroplasticity compared to the behavioral ones (Andrés, Parmentier, & Escera, 2006).

## Conclusion

The rise of DS prevalence sets a significant challenge in developing innovative health care interventions that provide the best quality of care. Neurocognitive treatments can augment the functional and cognitive abilities of PwDS and thus allow for a more independent, productive, and fulfilling daily life. Our results, based on brain imaging, connectivity, and graph-theory analysis, emphasize the significance of introducing stimulation and adaptable challenges in the environment of PwDS. We provide evidence that triggering adaptational neuroplasticity in the DS brain provokes the emergence of a less random, more hierarchical, and flexible network, able to integrate and segregate information more efficiently. Designing focalized neurobehavioral interventions can aid in the development of a balanced and stable functional DS phenotype in terms of SW characteristics.

### Materials and Methods Subjects

The study’s pool consisted of 12 subjects with DS (age: 29±11, 6 females). Participants were recruited from a variety of local organizations (please see Acknowledgements). The training procedure took place in the Thessaloniki Active and Healthy Ageing Living Lab (Thess-AHALL) (Konstantinidis, Billis, Bratsas, Siountas, & Bamidis, 2016), and the premises of the Greek Association of Down Syndrome. This study is part of the dsLLM clinical trial, registered with ClinicalTrials.gov, with identifier code NCT04390321. The study protocol was approved by the Bioethics Committee of the Medical School of the Aristotle University of Thessaloniki and was conducted per the Helsinki Declaration of Human Rights. The participants’ legal guardians signed written informed consent before their inclusion in the study.

### LLM Care Intervention

The intervention protocol consisted of physical and cognitive training (LLM Care (Bamidis et al., 2015), http://www.llmcare.gr/en). The study protocol was further developed to improve the quality of life and aid in the development of independent living skills in PwDS (Bamidis, Konstantinidis, Billis, & Siountas, 2017; Romanopoulou, Zilidou, Savvidis, Chatzisevastou-Loukidou, & Bamidis, 2018). LLM Care also aims to further improve brain functionality (Bamidis et al., 2015). All training sessions were computerized, center-based, and conducted under supervision. The sequence of training methods was pseudo-randomized and counterbalanced. The details of each training intervention have been previously described in detail (Billis et al., 2010; Smith et al., 2009) and are summarized in Figure 4.

**Figure 4.**
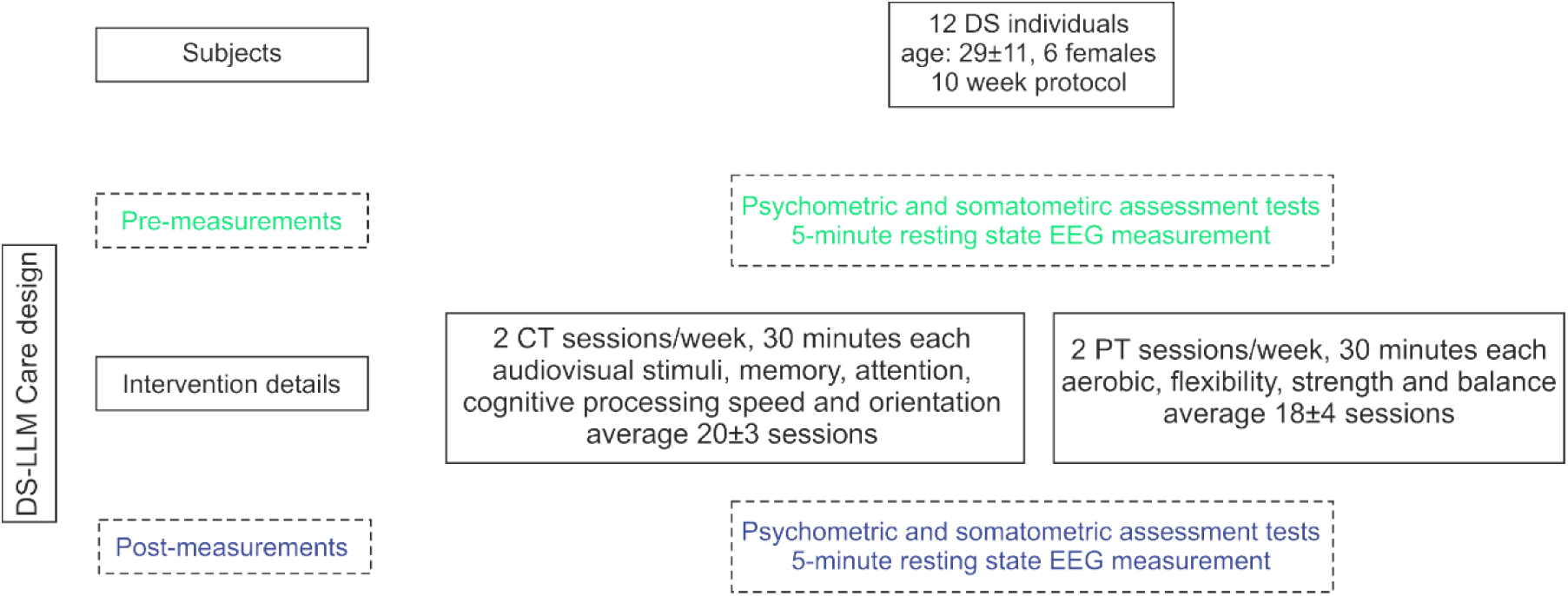
DS-LLM Care design and flow of participants with DS.

### Cognitive training

The CT component of LLM Care uses the BrainHQ software (Posit Science Corporation, San Francisco, CA, USA), an online interactive environment in the Greek language (Romanopoulou, Zilidou, & Bamidis., 2017). It consists of six categories (29 exercises in total), with customizable difficulty levels, utilizing audiovisual stimuli. CT targets memory, attention, cognitive-processing speed, navigation, and people skills. This regime was selected since patients with DS exhibit deficiencies in these processes. CT was conducted for half an hour, with a frequency of 2 days per week for 10 weeks. In every CT session, the participants were required to complete at least one task from each category. During the training, participants were urged to complete as many exercises as they could from each category.

### Physical training

The PT component of LLM Care is based on the WebFitForAll protocol (Billis et al., 2010; Konstantinidis, Bamparopoulos, & Bamidis, 2016) adequately adjusted to the needs and capacity of PwDS. It utilizes motion sensor devices (i.e., Kinect). Games and physical exercise are combined, providing a pleasant experience throughout the training. PT sessions lasted for half an hour and were conducted with the same frequency as CT. The training consists of aerobic (cycling, in-place-hiking), flexibility (stretching), strength (resistance, weightlifting), and balance (static, dynamic) exercises. The warm-up and cool-down routines (5-minutes duration) signify the start and completion of every session, respectively. During aerobic exercises, the participants entered a virtual environment, set up in Google maps, and explored cities and landscapes. Upon correct completion of the flexibility and strength exercises, the trainees were progressively rewarded with an array of pleasing images. The scope of balance exercises was to move their bodies either horizontally or vertically, which was achieved through games.

### Psychometric and somatometric assessments

The participants’ cognitive and physical capacity was assessed before and after the intervention. The psychometric evaluation consisted of a set of neurocognitive tests that measure memory, attention, concentration (WISC-III: Digits Span) (Woolger, 2001), verbal and non-verbal mental capabilities (Raven) (John & Raven, 2003), processing speed (WISC-III: Digits Span and Picture Arrangement), problem-solving, visuospatial processing, organization skills (WISC-III: Block Design, Picture Arrangement, Mazes), social intelligence (WISC-III: Picture Arrangement), and identification of emotions (Reading the mind in the eyes (Baron-Cohen, Jolliffe, Mortimore, & Robertson, 1997; Baron-Cohen, Wheelwright, Hill, Raste, & Plumb, 2001), and a variation: Reading the mind in the face (emotion recognition from video)).

The somatometric evaluation included the Short Physical Performance Battery (SPPB) (Guralnik et al., 1994), 10 Meter Walk (Bohannon, 1997), Back Scratch (Jones & Rikli, 2002), Sit and Reach (Wells & Dillon, 1952), Arm Curl (Jones & Rikli, 2002), Four Square Step (FSST) (Whitney, Marchetti, Morris, & Sparto, 2007), Stork Balance (for both legs) (Johnson & Jack K. Nelson, 1979), Timed Up and Go (Shumway-Cook, Brauer, & Woollacott, 2000) tests, and Body Mass Index (BMI). These tests appraise functioning mobility, flexibility (in specific areas), dynamic stability, strength, and static and dynamic balance.

### EEG recording

Pre- and post-intervention resting-state EEG activity was recorded for 5 minutes, using a high-density Nihon-Kohden EEG device (128 active scalp electrodes) at a sampling rate of 1000Hz. The EEG recordings were performed in an electrically shielded, sound, and light attenuated booth. The electrode impedances were lower than 10 kΩ. The participants were instructed to remain in a resting position while keeping their eyes open. Eyes-closed EEGs were not measured due to the limited capacity of the participants with DS to stay relaxed with their eyes closed.

### EEG data analysis Pre-processing

The raw EEG data were visually inspected, and bad channels were interpolated while any eye-movement related artifacts (blinks and horizontal movement) were corrected through adaptive artifact correction (Ille, Berg, & Scherg, 2002) using the Brain Electrical Source Analysis software (BESA research, version 6, Megis Software, Heidelberg, Germany) (Figure 5, blue section). The artifact-corrected data from each measurement were imported into the Fieldtrip Matlab toolbox (Oostenveld, Fries, Maris, & Schoffelen, 2011) for additional processing (Figure 5, blue section). The signals were denoised by filtering the data (0.53 Hz high-pass IIR filter, 48-52 Hz notch IIR filter, 97 Hz low-pass IIR filter). The filtered signals were analyzed into independent components (Lee, Girolami, & Sejnowski, 1999), and the artifactual components were rejected. After signal reconstruction, the data was, once again, visually inspected for any remaining artifacts. Fifteen segments (4 seconds each) from each artifact-free EEG recording were randomly selected for additional processing.

**Figure 5.**
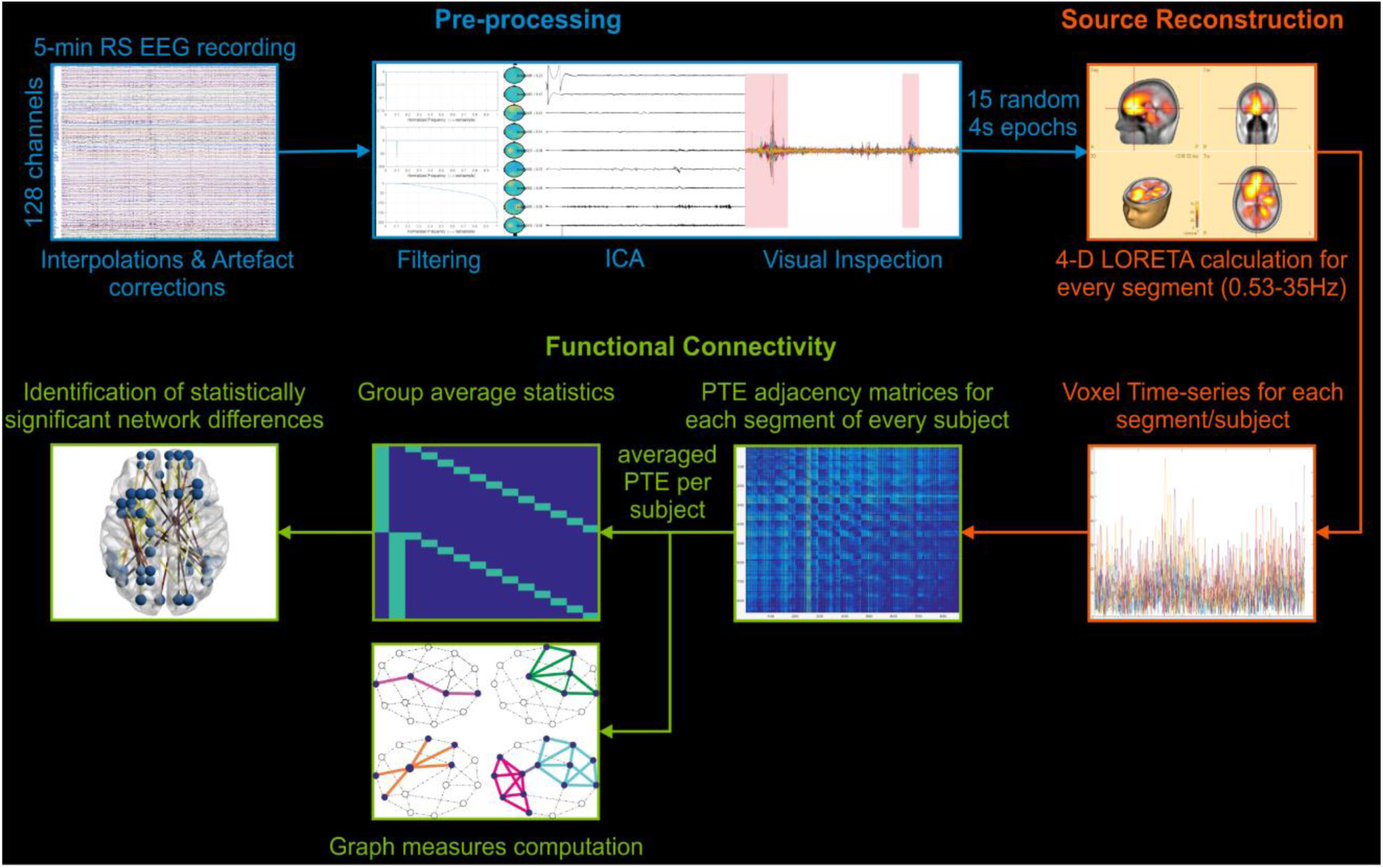
EEG data analysis schematic. **Pre-processing** (blue): EEG data were interpolated and artifact corrected, visually inspected, high-pass, bandpass, and low-pass filtered. Independent component analysis (ICA), as well as visual inspection, were used to reject artifactual data. 15 segments of 4-s were randomly selected. **Source reconstruction** (orange): The data were processed within 0.53-35 Hz frequency range, source reconstructed (4-D LORETA, for all time points), and a previously used 863-node atlas (Paraskevopoulos et al., 2015) was applied to extract the time-series of every voxel from every segment per subject, which were used to estimate 1000 surrogate time-series per subject/group. **Functional connectivity** (green): Functional connectivity was computed for every segment of every subject, using the phase transfer entropy metric (PTE), and the 15 matrices of every subject were averaged into one. PTE networks were estimated for every surrogate time-series of each subject to create a null-model for the rejection of noise in the real data. A network science approach was taken for the computation of graph measures per subject. Group average statistics were calculated to identify the statistically significant differences between post- and pre-intervention networks.

### Source reconstruction

Each subject’s segments (15 segments, 4000 samples, 4 seconds duration each) were imported into BESA (Figure 5, orange section). The current density reconstructions (CDR) were estimated for each sample point, solving the inverse problem using LORETA in the 0.5-35 Hz frequency range. LORETA (Pascual-Marqui et al., 1994) was utilized, as it does not require the *a priori* declaration of the number of sources and is suitable for whole cortex analysis. The CDRs were exported as four-dimensional images (4-D) in the Analyze format (keeping all sampling points), which were, in turn, imported into Matlab. A cortex mask was superimposed on the images. The mask includes only grey matter and excludes the subcortex, the brainstem, and cerebellum, to limit the source space (Paraskevopoulos, Kraneburg, Herholz, Bamidis, & Pantev, 2015). The source space consisted of 863 voxels.

### Functional connectivity

After extracting the time-series of each voxel from the 4-D images, they were used to compute the Phase Transfer Entropy (PTE) (Lobier, Siebenhühner, Palva, & Palva, 2014) (Figure 5, green section). The computation resulted in 863×863 adjacency matrices. The metric is calculated independently for every pair of voxels in each segment. Each voxel represented a node of the brain network, with the node’s coordinates corresponding to the center of each voxel. PTE was selected because its results are not based on a specific data model since its computation is reliant on non-linear probability distributions. Therefore, it allows the detection of higher-order relations in the phase information flow and renders the measure resistant to source leakage (Lobier et al., 2014). The algorithm applies the Hilbert transform to estimate the phases of each signal. It determines the number of bins by utilizing the Scott methodology (Scott, 1992), resulting in 37 bins on average (13 samples per bin). Each subject’s adjacency matrices were averaged, resulting in two sets of 12 adjacency matrices (Figure 5, green section).

When computing network science indices based on information theory, one should take into account that the derived data contain noise that may mask the “real” result. This potential limitation was dealt with by following a methodology proposed by Timme et al. (Timme et al., 2016). Thus, subject-level null-distributions were computed and then compared with the real PTEs. 2000 surrogate time-series were estimated for each subject (1000 for the pre-intervention measurements and 1000 for the post-intervention ones) via phase randomization, i.e., calculating the Fourier transform of the real times-series, randomizing their phase, inverting the transform, thus resulting in jittered time-series (Timme et al., 2016). PTE networks were estimated from the shifted time-series and then compared edge by edge to the subject-PTE-networks, where any real-edge that had lesser weight than more than 50 (5% significance level) surrogate-edges was considered non-significant and removed from the final PTE network (Timme et al., 2016). To ensure that small differences between the surrogate and real data did not affect the edge-significance test, the weight difference between them was estimated, and differences less than e-03 were ignored. The comparison of the surrogate PTE distributions to the PTE data resulted in networks consisting of 863 nodes and 184720 edges on average for the post-intervention networks and 123980 edges on average for the pre-intervention networks.

### Graph measures computation

Using the Brain Connectivity Toolbox (Rubinov & Sporns, 2010), GE, TS (a variation of global CC), CPL, CC, BC, as well as community detection, and the measure of SW (σ) were computed for each participant. In a later step, the centrality Matlab function, which measures node importance, was used to calculate the node degree centrality (DC) of the nodes showcasing significant shits in the post-intervention network. The density of each graph was estimated by averaging the weights of each graph and multiplying the average by the maximum weight of the network.

These graph measures were chosen to examine the influence of our intervention on the characteristics of the DS brain and its functional organization. To that aim, GE and CPL were utilized to index the effects on integration (Rubinov & Sporns, 2010), TS and CC to measure the impact on segregation (Rubinov & Sporns, 2010), BC and DC to check for shifts in the importance of nodes to the flow of information, and community detection to characterize the hierarchical organization.

GE is the average inverse shortest path length of the network (Latora & Marchiori, 2001) and quantifies the efficacy of information transference and its assimilation in the network (Rubinov & Sporns, 2010). CPL is the average shortest path length of the edges connecting the nodes of the network (Watts & Strogatz, 1998), is influenced by longer paths, and characterizes the robustness of the network (Rubinov & Sporns, 2010). TS is the ratio of closed triplets to the maximum number of triplets (open and closed) (Newman, 2003), while CC of a node is the ratio of its connected neighbors to the maximum number of possible connections (Watts & Strogatz, 1998). These two measures reflect the clustering organization of the DS brain network on a global and local level, respectively (Rubinov & Sporns, 2010). BC corresponds to the fraction of shortest paths that pass through a node (Freeman, 1978) and is a measure of centrality, measuring the importance of a node in the information flow (Rubinov & Sporns, 2010), and DC estimates the number of links connected to a specific node, i.e., it measures if a node exhibits higher or lower connectivity to other nodes. To characterize the DS network’s hierarchical organization, the number of communities that constitute the pre and post DS networks was estimated. Community detection was achieved through the implementation of a multi-iterative generalization of the Louvain community detection algorithm that optimizes modularity (Blondel, Guillaume, Lambiotte, & Lefebvre, 2008). Following the methodology of Akiki & Abdallah (Akiki & Abdallah, 2019), the detection was applied recursively to pinpoint the multi-scale, hierarchical community organization. A single-level detection was performed for 100 iterations for every subject, and from them, a group-representing partition was selected using the partition similarity measure of normalized mutual information. This partition was then used as a base, to extract subgraphs from each community, which were then fed into the algorithm for the next level of community detections. This process was repeated until no new communities were detected. The measure quantifying a network’s small-worldness is sigma (σ). It is based on the ratio of the comparison of a network’s CC and CPL to that of random networks (Bolaños, Bernat, He, & Aviyente, 2013; Humphries & Gurney, 2008) with the network characterized as SW when *σ>1.* To estimate the SW measure, we generated 100 random networks from each subject’s pruned adjacency matrix while preserving the out-degree, with each edge being reshuffled at least 100 times, and calculated the σ in reference to each random network. Finally, we averaged the σ-values per subject. These random networks were also used to create a personalized null-distribution for each subject to mitigate the effect of density on graph measures when reporting individual results. For a summary of the graph measures used in the analysis, please refer to the Supplementary Material (Supplementary Table 1). The comparison of our pre- and post-intervention network in relation to the exhibition of random or regular characteristics are discussed on a qualitative level, i.e., whether they show characteristics exemplary for a random organization (small CPL, low CC). These random characteristics are present in many random topologies, and hence we have selected the most simplified random organization, the Erdös–Rényi. The degree distribution of the DS networks is taken into consideration when this comparison is quantified (i.e., estimation of SW), where a generative random model that preserves the degree distribution (Newman, 2002) of the DS networks is used.

### Statistical analysis

The pre- and post-intervention somatometric and psychometric assessment scores were compared with the use of non-parametric Wilcoxon tests and paired t-tests. The statistical comparisons were performed using the IBM SPSS 25.0 software. Wilcoxon tests were performed on the psychometric battery tests and somatometric tests with a discrete-values score, while paired t-tests were applied on the remaining somatometric assessment tests.

The Network Based Statistics (NBS) (Zalesky et al., 2010) MATLAB toolbox was employed to estimate the statistically significant differences between the whole-head network of the pre- and post-intervention connectivity networks of our subjects. A paired samples t-test corrected for 5000 random comparisons via the non-parametric NBS method (Zalesky et al., 2010) was performed. The permutation model of the NBS toolbox randomly exchanges the group label of each subject at every permutation level and then recalculates the statistic of interest applying the selected threshold. The maximal component size is derived from each permutation and utilized to create an empirical estimate of the null-distribution of maximal component size (Zalesky et al., 2010). For further information on this process, please refer to the following work (Hayasaka & Nichols, 2004; Nichols & Holmes, 2002). Significant differences between the two time-points were visualized as weighted graphs through the BrainNet Viewer (Xia, Wang, & He, 2013) toolbox (Figure 1). DC was estimated from the outcome of this comparison, and the results were depicted in the same graph.

For the graph measures, Analysis of Covariance (ANCOVA) was used, where each measure serves as the dependent variable with density as the covariate because density seriously affects the values of the other measures. For local measures (i.e., CC and BC), ANCOVA and FDR correction were performed for each of the 863 nodes and p-values for Type I errors, respectively. Results below the 5 percent threshold were considered significant.

## Data Availability

All data underlying the findings described in the manuscript will be fully available without restriction upon request

## Acknowledgments

This study is an extension of the European CIP-ICTPSP. 2008.1.4 Long Lasting Memories (LLM) project (Project no. 238904) (http://www.longlastingmemories.eu/). The authors would like to acknowledge the support (i.e. high-performance computing infrastructure and resources) provided by the IT Center of the Aristotle University of Thessaloniki (AUTH) throughout the progress of this research work. The authors would like to thank all the participants, their families, and caregivers, the Down Syndrome Association of Greece, Spring Children Mixed Living Center – Daily Care Activity & Training for Disabled People, The Down Syndrome Association of Serres, Center for the Rehabilitation of Social Support & Creative Employment of People with Disabilities, Child Care Center in Ptolemaida, Centre For Persons With Learning Difficulties - Zoodohos Pigi, Center of Creative Activities - Municipality of Neapoli and Sykies, Special laboratory for vocational education and training in Alexandria, Center for the Rehabilitation of Social Support & Creative Employment of People with disabilities "O Sotir", Center of Creative Activities for People with Disabilities-Municipality of Neapoli and Sykes. Also, the authors would like to thank a list of people assisting in the trial facilitation, as follows: Dimitris Bamidis, Foteini Dolianiti, Ioanna Dratsiou, Sotiria Gilou, Maria Karagianni, Katerina Katsouli, Eleni Manthou, Maria Metaxa, Annita Varella (in alphabetical order), as well as, students from the Dept of Psychology, The University of Sheffield International Faculty, City College (Greece) who were practicing under the supervision of Dr. Anna Vivas and Dr. Elissavet Chrysochoou who were also advising the authoring team on the psychological aspects of the whole endeavor. Last but not least, the authors would like to thank Emeritus Professor of Pediatrics and Genetics Dr. Charikleia Chatzisevastou-Loukidou for advising the team on all medical aspects of the project and for linking the team with many of the pilot sites.

