## Supplementary Material for "Computerized physical and cognitive training improves the functional architecture of the brain in adults with Down Syndrome: a network science EEG study"

* Joined first authorship

** Joined senior authorship

### Supplementary Information

The following steps were taken to get a measure of how much of the entropy of the receiver is explained by information flowing from the sender. Values of normalized PTE (dPTE) range between 0 and 1, with weights of node-pairs xy in the interval [0, 0.5) indicating that information flows from x to y, weights in the interval (0.5,1] showcase the opposite direction of information, while weights with a value of 0.5 indicate the equal participation of x and y (Hillebrand et al., 2016). We have averaged the dPTE adjacency matrices per group and then visualized the edges with weights $\leq 0.5$ to depict the participation of the receiving node in the information flow i.e., the higher the weight of an edge the more the receiver participates in the exchange of information. The pre-intervention network seems to have a stronger looping effect in the information flow, as certain receiver nodes participate almost reciprocatively in the exchange of information within the pair. This effect appears to have decreased in the post DS network but an increased amount of receiver nodes seems more engaged in the communication within the network.

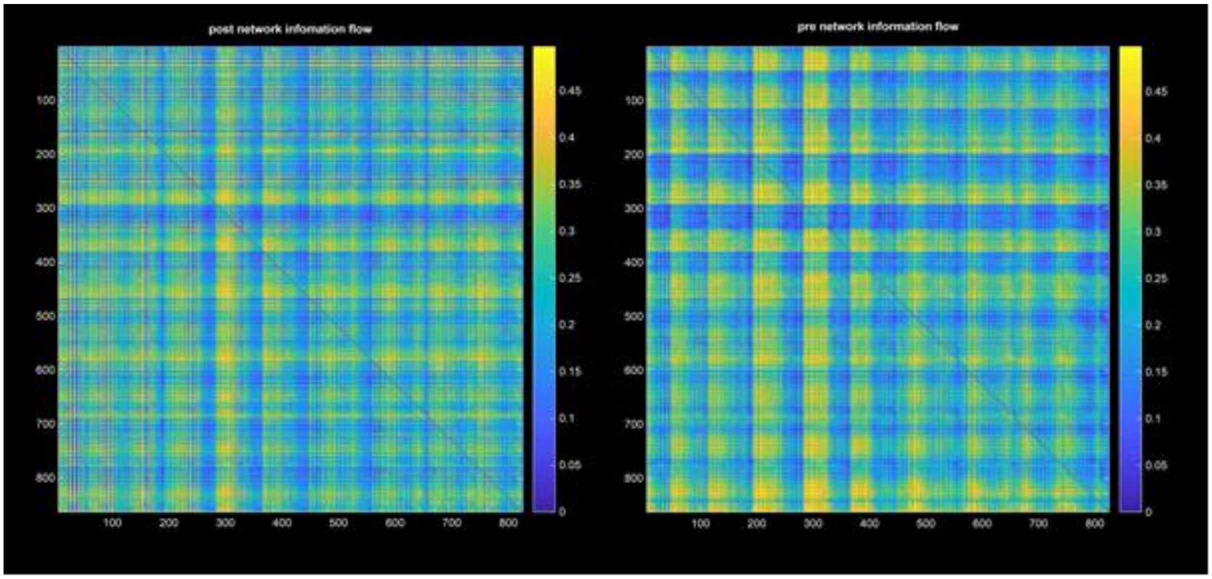


**Supplementary Figure 1**. Depiction of the engagement of receiver nodes in the exchange of information within the network. Values of normalized PTE (dPTE) range between 0 and 1, with weights of node-pairs xy in the interval [0, 0.5) indicating that information flows from x to y, and weights in the interval (0.5,1] showcase the opposite direction of information, while weights with a value of 0.5 indicate the equal participation of x and y (Hillebrand et al., 2016). The colormap depicts the engagement of nodes receiving information in the communication with their pair (*dPTE≤0.5*). The greater the value, the more engaged the receiver node is in the exchange of information.

**Supplementary Table 1.** Summary of the graph measures used in the analysis

<table>
<tr><th colspan="3">Graph Measures</th><th>Definition</th><th>Relation to other measures</th><th>Use</th></tr>
<tr><td rowspan="3">Global Measures</td><td rowspan="2">Measures of integration</td><td>Global Efficiency</td><td>The average inverse shortest path length. Efficacy of information transference.</td><td>Inverse relation to the characteristic path length</td><td>Characterization of shifts in integration and cost-efficiency of the network</td></tr>
<tr><td>Characteristic Path length</td><td>The average shortest path length between nodes of the network. Number of edges that need to be traversed for the communication between distant regions of the network.</td><td>Inverse relation to global efficiency<br>Related to the small-worldness measure</td><td>Characterization of the pre- and post-DS network organization</td></tr>
<tr><td rowspan="2">Measures of segregation</td><td>Transitivity</td><td>The ratio of closed triplets of nodes to the maximum number of triplets (open and closed).<br>Global clustering organization of the network.</td><td>Related to the local clustering coefficient (average clustering coefficient).<br>Related to the small-worldness measure.<br>Does not suffer from individual disproportionate clustering coefficient values</td><td>Characterization of the pre- and post-DS network organization, and shifts in the segregation of the network</td></tr>
<tr><td rowspan="2">Local Measures</td><td>Local Clustering Coefficient</td><td>The ratio of a node's connected neighbors to the maximum number of possible connections. Clustering organization in distinct regions that characterizes the robustness of a system.</td><td>Related to transitivity</td><td>Characterization of shifts in the robustness and fault tolerance of the DS network</td></tr>
<tr><td>Measures of centrality</td><td>Node Betweenness Centrality</td><td>The fraction of shortest paths that pass through a node.<br>Importance of a node in the information flow</td><td>Analogous relation to degree centrality</td><td>Characterization of the participation of nodes in the distribution of information</td></tr>
</table>

| | | | | | |
|---|---|---|---|---|---|
| | | Node Degree Centrality | Number of links connected to a node. The connectedness of a node to others. | Analogous relation to degree centrality | Computed only for the nodes of the post-DS network, which exhibit a significant increase in connectivity |
| | Network motifs | Communities | Detection of communities' organization by maximizing modularity. Characterizes the community distribution of a network. | | Characterization of the hierarchical organization of the pre- and post- DS networks |
| | | Small-Worldness | The ratio of global clustering coefficient and characteristic path length, normalized by the respective measures of a random network. Characterizes the organization of a network in relation to the threshold of small-worldness ($\sigma>1$ classifies a network as small-world). | Related to the characteristic path length and transitivity | Characterization of the pre- and post-DS network organization. |
